# Patient and Surgeon Willingness to Participate in a Randomized Trial of Surgery Versus Observation for Mild Cervical Spondylotic Myelopathy: A Cross-Sectional Survey Study

**DOI:** 10.64898/2026.08.18.26360719

**Authors:** Faraz Arkam, Xianqi Zeng, Eliana Goldstein, Jetan Badhiwala, Andrew K. Chan, Abby L. Cheng, Dean Chou, Matthew Colman, Zoher Ghogawala, Jakub Godzik, Michael P. Kelly, Thomas E. Mroz, Brian J. Neuman, Lindsay Orosz, Paul Park, Alpesh A. Patel, Eric A. Potts, John Rhee, Rick C. Sasso, Kenneth B. Schechtman, Michael P. Steinmetz, Grace X. Xiong, Salim Yakdan, Linying Zhang, Wilson Z. Ray, Mary C. Politi, Jacob K. Greenberg

## Abstract

**Objective:** Cervical spondylotic myelopathy (CSM) is the most common cause of nontraumatic spinal cord dysfunction in adults. For mild disease, guidelines recommend shared decision-making between surgery and structured rehabilitation, yet no comparative effectiveness study has reported outcomes in this population. Whether a randomized trial is feasible is unknown.

**Methods:** We conducted two cross-sectional surveys between December 2025 and July 2026: one of patients with surgeon-confirmed CSM of any severity, recruited from academic outpatient spine clinics, and one of practicing neurosurgical and orthopedic spine surgeons. Respondents rated willingness to participate in (1) a randomized trial of early surgery versus observation and (2) a prospective observational study in which treatment was patient selected. Responses of likely or very likely were classified as willing. Groups were compared using Fisher exact tests, designs within respondents using exact McNemar tests, and predictors using univariable logistic regression.

**Results:** Fifty-six patients and 52 surgeons completed the surveys. Patients were markedly less willing than surgeons to accept randomization (16 of 56, 28.6% versus 44 of 52, 84.6%; p < 0.001). Both groups accepted the observational design (41 of 56, 73.2% versus 51 of 52, 98.1%; p < 0.001), and 27 of 40 patients unwilling to be randomized were willing to enroll in an observational study (p < 0.001). Willingness to be randomized did not differ across mJOA severity (mild 29.2%, moderate 25.0%, severe 33.3%; p = 0.93). Among patients declining randomization, 85.2% cited a wish to retain control over treatment, whereas fear of surgery was cited by one respondent. Forty-five surgeons (86.5%) considered both surgery and observation reasonable, and preference was divided (46.2% favoring early surgery, 48.1% favoring initial observation).

**Conclusions:** Surgeon responses were consistent with equipoise, and most surgeons were willing to randomize, but most patients would decline random allocation, citing a wish to retain treatment choice rather than fear or distrust. A prospective observational study appears as the more feasible route to comparative evidence in mild CSM. Feasibility assessments restricted to clinicians may overestimate attainable accrual.

## INTRODUCTION

Cervical spondylotic myelopathy (CSM) is the most frequent cause of non-traumatic spinal cord dysfunction in adults worldwide,^1,2^ with a prevalence of up to 2%.^3^ Patients with CSM experience progressive gait impairment, loss of hand dexterity, sensory disturbance and, in advanced disease, bladder dysfunction.^1,4^ Quality-of-life scores in CSM are among the lowest reported for chronic disease.^5,6^

Surgical decompression is the mainstay of treatment for moderate to severe CSM and remains the only intervention supported by high-quality evidence.^4^ The 2017 AO Spine/Cervical Spine Research Society clinical practice guideline issued a strong recommendation that surgery be offered to patients with moderate (modified Japanese Orthopaedic Association [mJOA] (12–14) and severe (mJOA ≤ 11)) myelopathy.^7^ For these patients, the benefits of surgery outweigh its risks, and patients report positive post-operative improvement in quality of life.

However, for patients with mild symptoms, the same guideline recommends clinician-based or shared decision-making about whether to pursue surgical intervention or supervised, structured rehabilitation based on clinical equipoise between options. There is no comparative effectiveness study reporting outcomes between options in patients with mild disease.^7^ Estimates of both the benefit of surgery and the risk of nonoperative management remain poorly defined. For example, reported rates of neurological deterioration under nonoperative management range from 13.3% at one year in a cohort restricted to mild CSM to 57% over a mean of 2.5 years in another.^8–10^ Optimizing the application and timing of surgery in milder cases has accordingly been designated a top research priority by the AO Spine RECODE-DCM international multi-stakeholder priority-setting partnership.^11,12^

A randomized comparison of early (upfront) surgery against structured nonoperative care is one approach to resolve this uncertainty. However, no such trial has been completed, and its feasibility is unknown. Randomized trials in CSM have instead compared surgical techniques: the PCORI-funded Cervical Spondylotic Myelopathy Surgical (CSM-S) trial, compared ventral with dorsal surgical approaches in 163 patients.^13,14^ That trial included no nonoperative arm.

Whether such a comparison applies to mild disease has not been examined. Surgeon and patient treatment preferences are among the most frequently cited barriers to recruitment in surgical trials, affecting both attainable sample size and the generalizability of any result.^15^ Spine surgery offers a cautionary precedent: the Spine Patient Outcomes Research Trial (SPORT) enrolled concurrent randomized and observational cohorts precisely because treatment preference was anticipated, and its randomized cohorts experienced crossover so extensive that intention-to-treat analyses became difficult to interpret.^16–18^

To our knowledge, no study has assessed equipoise among surgeons treating mild CSM or the willingness of patients and surgeons to enroll in a trial of surgery versus observation. We therefore conducted separate cross-sectional surveys of patients with CSM and of practicing spine surgeons. Our objectives were to (1) compare patients’ willingness to participate in a randomized controlled trial versus a prospective observational study of early surgery versus observation for mild CSM; (2) compare surgeons’ willingness to enroll patients in the two study designs; (3) identify patient-level factors, including myelopathy severity, associated with willingness to participate in each design; and (4) assess whether clinical equipoise exists among surgeons regarding early surgery versus observation for mild CSM.

## METHODS

### Study design and reporting

We conducted two separate cross-sectional surveys, one of patients with CSM and one of practicing spine surgeons. The study is reported in accordance with the Consensus-Based Checklist for Reporting of Survey Studies (CROSS) and with the Checklist for Reporting Results of Internet E-Surveys (CHERRIES).^19,20^ Both completed checklists are provided as Supplementary Files 1 and 2.

The study was approved as exempt research by the Institutional Review Board at Washington University in St. Louis under the approval ID #202511100. Patients and surgeons reviewed a consent information sheet prior to completing the survey. No compensation was offered to patients or surgeons.

### Participants and recruitment

Patients were eligible if they were adults with a diagnosis of CSM confirmed by an attending spine surgeon based on clinical findings consistent with myelopathy and radiographic evidence of cervical spinal cord compression. Patients were invited by their treating surgeon when the choice between surgery and nonoperative management was under active consideration or had recently been made, which is the point at which enrollment into a prospective study would occur. Severity was classified afterwards from the mJOA score collected in the survey, and all completed responses were analyzed regardless of severity. Patients were recruited from academic outpatient spine clinics between December 2025 and July 2026. They received the survey link through flyers or online posts, a QR code scanned during a clinic visit, or encrypted email sent with their consent after being contacted during a routine visit or by phone.

Surgeons were eligible if they were practicing neurosurgical or orthopedic spine surgeons involved in the management of CSM. Surgeons were identified through the investigators’ professional networks and invited by email over the same period, using a snowball approach in which recipients were asked to forward the link to eligible colleagues.

76 patients who accessed the survey, 63 (83%) provided consent and 56 (89% of those consenting) completed the survey. The survey was emailed directly to 84 surgeons. 65 surgeons who accessed the survey, 62 (95%) provided consent and 52 (84% of those consenting) completed it. Because forwarded invitations could not be tracked, the total number of surgeons reached is unknown; relative to direct invitations alone, the response rate was at most 62%.

### Survey instruments

Both instruments were developed by the study team, which included CSM clinicians, comparative-effectiveness researchers and survey-design experts, informed by clinical literature and by clinical experience in the management of CSM. Before distribution, members of the study team reviewed the instruments for clarity, relevance, and content. The full instruments are provided as Supplementary Files 3 and 4.

The patient instrument collected demographic characteristics, symptom duration, prior spine surgery at any spinal level, current position in the treatment pathway, and the self-administered modified Japanese Orthopaedic Association (mJOA) score, which assesses upper-extremity motor function, lower-extremity motor function, upper-extremity sensation, and bladder function. Respondents then rated, on five-point Likert scales, how likely they would be to participate in (a) a randomized trial allocating patients to early (upfront) surgery or to observation, and (b) a prospective observational study in which they selected their own treatment. Patients indicating they were unlikely or very unlikely to accept randomization were asked to identify the reasons underlying that reluctance from a fixed list adapted from a prior prospective preference assessment of surgical versus nonoperative care, selecting all that applied.^21^ All patients were additionally asked which changes in their condition would prompt a switch from observation to surgery (select all that apply), and which outcome mattered most in defining recovery.

The surgeon instrument collected demographic and practice characteristics, willingness to enroll patients in each of the two study designs, and four Likert-scaled items assessing equipoise regarding the management of mild CSM. Surgeons unwilling to randomize were asked to select their reasons from a fixed list developed by the study team. Current practice patterns were also assessed, including the estimated proportion of patients undergoing early surgery and the frequency of recommending structured physical therapy.

### Survey administration

Surveys were administered electronically using REDCap electronic data capture tools.^22,23^ Responses were anonymous and no identifiers were collected. The patient instrument comprised of 20 items presented across 4 sections and the surgeon instrument 17 items across 4 sections. Adaptive questioning was applied to the reasons-for-reluctance item in each instrument: only patients indicating they were unlikely or very unlikely to accept randomization were shown the question regarding reasons for reluctance. Respondents were able to review and revise their responses before submission. Respondents who did not complete the survey were excluded; for individual items, percentages are reported among those who answered.

### Definitions and outcome measures

#### Willingness

The primary outcome for each respondent group was willingness to participate in each study design. Responses of “likely” or “very likely” were categorized as willing and all other responses as not willing. Respondents answering “unlikely” or “very unlikely” are referred to as unwilling, and those answering “neither likely nor unlikely” as neutral.

#### Myelopathy severity

Myelopathy severity was classified using the mJOA as severe (≤ 11), moderate (12–14) or mild (≥ 15).^7,24^

### Statistical analysis

Continuous variables are summarized as mean ± standard deviation and categorical variables as counts with percentages. Proportions are reported with 95% Wilson score confidence intervals. Comparisons of willingness between patients and surgeons, who constituted independent samples, were made using Fisher exact tests. Within-respondent comparisons of willingness between the two study designs were made using exact McNemar tests. Comparisons of willingness across mJOA severity categories were made using exact tests of independence, as expected cell counts were below five.

Factors associated with willingness were examined using univariable logistic regression, performed for each candidate predictor (age, sex, education, household income, symptom duration, prior spine surgery, mJOA severity category, and current position in the treatment pathway). Current position in the treatment pathway was categorized as actively deciding between surgery and observation versus all other responses.

Two-sided p values below 0.05 were considered statistically significant. No adjustment was made for multiple comparisons; analyses beyond the primary comparison of willingness between study designs are therefore regarded as exploratory and hypothesis-generating. Analyses were performed using R version 4.4.2. Univariable logistic regression was performed using base R (stats).

## RESULTS

### Respondent characteristics

Fifty-six patients with CSM and fifty-two practicing spine surgeons completed the surveys. Baseline characteristics are presented in Table 1 (patients) and Table 2 (surgeons).

Patients had a mean age of 64.1 ± 10.3 years, 33 (58.9%) were female and 50 (89.3%) identified as White. Symptom duration exceeded two years in 27 patients (48.2%) and 18 (32.1%) had undergone prior spine surgery. Respondents were distributed across the treatment pathway: 20 (35.7%) were actively deciding between surgery and observation, 14 (25.0%) were planning to undergo surgery, 14 (25.0%) were observing symptoms without plans for surgery, 4 (7.1%) had not yet discussed treatment options with a physician, and 4 (7.1%) had already undergone surgery.

Twenty-four patients (42.9%) had mild (mJOA ≥ 15), 20 (35.7%) moderate (12–14) and 12 (21.4%) severe (≤ 11) myelopathy.

Surgeons had a mean age of 46.4 ± 9.7 years and 48 (92.3%) were male. Primary specialty training was neurosurgery in 24 (46.2%), orthopedic spine surgery in 23 (44.2%) and dual training in 5 (9.6%). Forty-three surgeons (82.7%) practiced in an academic setting, and 43 (82.7%) reported treating more than 25 patients with mild CSM annually, of whom 22 (42.3%) treated more than 50 patients.

### Willingness to participate, by study design and respondent group

Patients were significantly less willing than surgeons to participate in a randomized trial: 16 of 56 patients (28.6%) compared with 44 of 52 surgeons (84.6%), an absolute difference of 56.0 percentage points (Fisher exact p < 0.001). Willingness to participate in the observational design was high in both groups but remained significantly greater among surgeons: 41 of 56 patients (73.2%) versus 51 of 52 surgeons (98.1%) (Fisher exact p < 0.001) (Figure 1).

**Figure 1.**
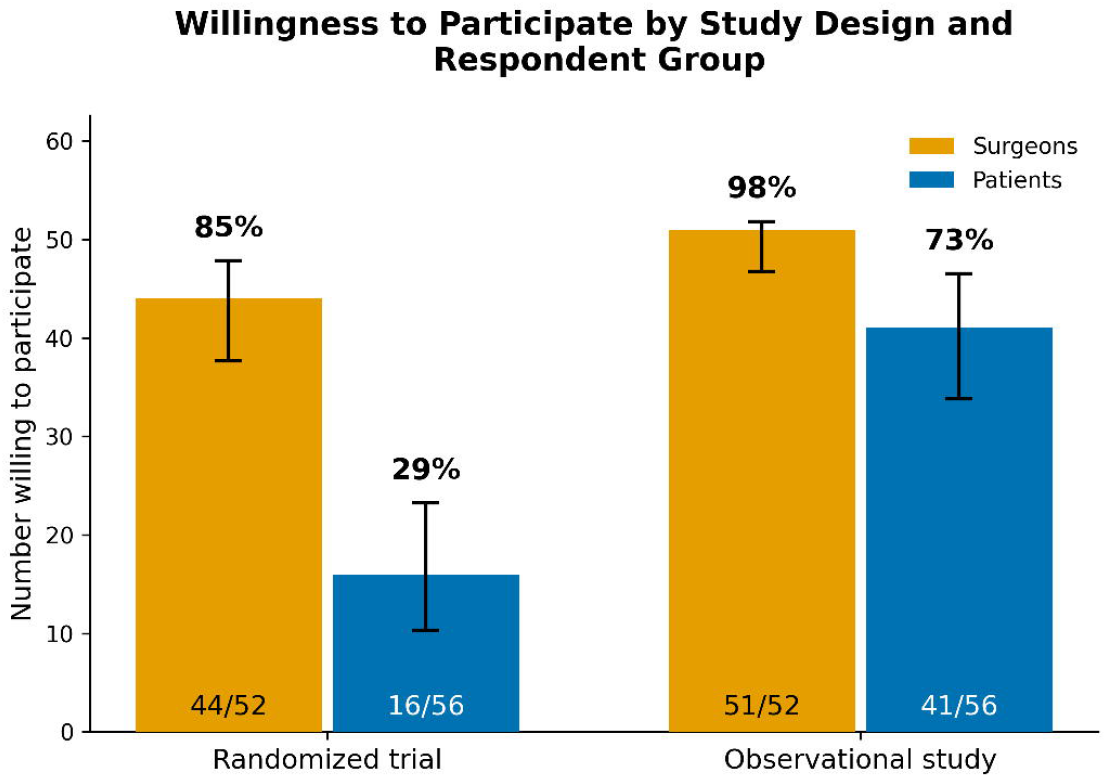
Willingness to participate by study design and respondent group.

Within each respondent group, willingness differed significantly between the two designs. Among patients, 27 of 40 who were not willing to be randomized were willing to join the observational study, whereas only 2 showed the reverse pattern (McNemar exact p < 0.001). All 8 surgeons not willing to randomize their patients were willing to enroll them in an observational study, and the single surgeon not willing to participate in the observational study was willing to randomize (McNemar exact p = 0.039).

### Consistency of findings across disease severity

Because the clinical question of surgery versus observation is most contested in mild disease, patient willingness was examined across mJOA severity categories to determine whether the findings were confined to any one stratum (Figure 2). Willingness to participate in a randomized trial did not differ by severity: 7 of 24 patients with mild disease (29.2%), 5 of 20 with moderate disease (25.0%) and 4 of 12 with severe disease (33.3%) were willing to be randomized (exact p = 0.93). Willingness to participate in the observational study also did not differ significantly by severity (mild 79.2%, moderate 75.0%, severe 58.3%; exact p = 0.46).

**Figure 2.**
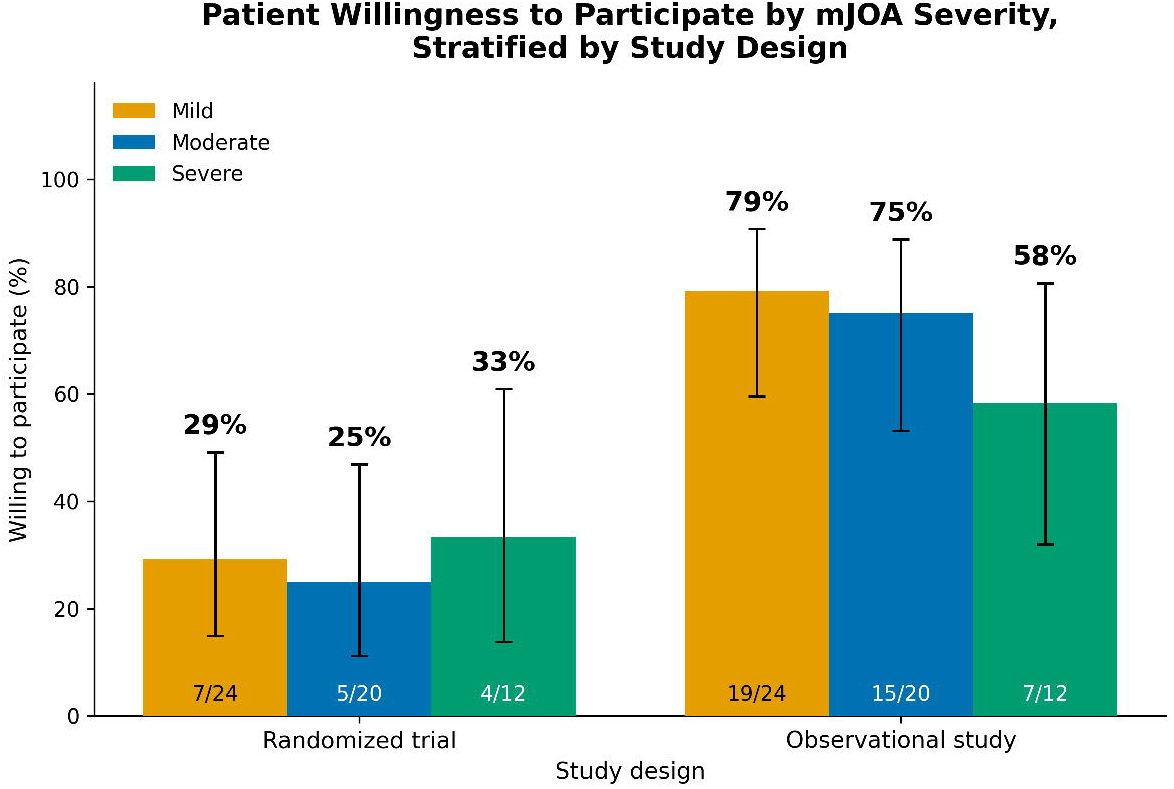
Patient willingness to participate by study design, stratified by mJOA severity.

### Reasons for reluctance to accept randomization

Patients who indicated they were unlikely or very unlikely to accept randomization (n = 27) were asked to identify the reasons underlying that reluctance. The most frequently endorsed reason was a wish to maintain control over treatment decisions (23 of 27, 85.2%), followed by fear that symptoms would worsen if surgery were delayed (8 of 27, 29.6%) and distrust of randomization as a method of treatment allocation (3 of 27, 11.1%). Fear of surgery itself was endorsed by a single respondent (1 of 27, 3.7%).

Eight surgeons expressed reluctance to participate in a randomized trial. The most frequently endorsed reasons were the inability to individualize treatment for each patient and ethical concerns regarding delayed surgery for patients allocated to observation. Given the small number of respondents these are reported descriptively and no statistical comparison was made.

### Factors associated with willingness to participate

Sixteen patients were willing to participate in a randomized trial. In univariable logistic regression, household income was inversely associated with willingness (OR 0.61 per one-level increase, 95% CI 0.39–0.92, p = 0.021), and patients currently deciding between surgery and observation were more likely to be willing (OR 3.39, 95% CI 1.03–11.78, p = 0.047), although the lower confidence limit approached 1. No other variable reached significance, including mJOA severity, symptom duration, sex, prior spine surgery, education or age (Table 3).

For willingness to participate in the observational study, female sex (OR 6.65, 95% CI 1.76– 25.07, p = 0.005) and currently deciding between surgery and observation (OR 12.09, 95% CI 1.45–100.69, p = 0.021) were significantly associated. Household income was not associated with willingness to participate in the observational design (OR 1.10, 95% CI 0.74–1.64, p = 0.64) (Table 4).

### Surgeon attitudes and equipoise

Surgeon attitudes toward the management of mild CSM are presented in Table 5. Forty-five of 52 surgeons (86.5%) agreed or strongly agreed that both surgery and observation are reasonable management options. Surgeon opinions regarding their individual preferred initial approach were near-evenly divided: 24 of 52 (46.2%) thought that early surgery leads to better long-term neurological outcomes, and 25 of 52 (48.1%) thought that an initial 6-to 12-month period of observation is more appropriate. More than 80% agreed that further comparative-effectiveness research is needed. Attitudes did not differ significantly by primary specialty training on any of the four items.

### Surgeon practice patterns

Surgeons reported a broad spectrum of practice with respect to early surgery for mild CSM (n = 52): 14 (26.9%) estimated that fewer than 25% of their patients with mild CSM undergo surgery within six months of presentation, 21 (40.4%) estimated 26–50%, 16 (30.8%) estimated 51–75%, and 1 (1.9%) estimated 76–100%. For patients managed nonoperatively, 35 of 52 surgeons (67.3%) reported recommending structured physical therapy often or always, and a further 11 (21.2%) reported recommending it sometimes.

### Patient-reported factors prompting a switch to surgery, and outcome priorities

Patients were asked which symptom or functional changes would prompt a switch from nonoperative management to surgery, selecting all that applied. Worsening neck or arm pain was selected most frequently (36 of 56, 64.3%), followed by decline in balance or walking ability (30 of 56, 53.6%), worsening hand function (25 of 56, 44.6%) and decline on clinician-administered tests such as walking speed (15 of 56, 26.8%).

When asked which outcome mattered most in defining recovery, 41 patients (73.2%) selected overall quality of life and physical function, 8 (14.3%) selected specific daily activities and 7 (12.5%) selected pain.

## DISCUSSION

In separate surveys of patients with CSM and of the surgeons who treat them, willingness to participate in a randomized trial of early surgery versus observation differed markedly between the two groups. Few (29%) patients indicated willingness to accept random allocation, compared with 85% of surgeons. Both groups, however, readily accepted a prospective observational design in which treatment was selected by the patient, and two-thirds of the patients who declined randomization remained willing to participate in such a study. Reluctance to be randomized was driven overwhelmingly by a wish to retain control over the treatment decision rather than by fear of surgery or distrust of research. Surgeon responses were consistent with the equipoise that existing guidelines describe.

This asymmetry between patient and surgeon willingness to be part of a randomized trial of treatment for CSM may carry an implication that extends beyond CSM: feasibility assessments conducted among investigators alone may systematically overestimate attainable accrual into randomized trials. This pattern is consistent with the broader surgical trials literature, in which a preference for choosing one’s own treatment is among the most common reasons for declining randomization, and contrasts with CSM-S, in which patients were randomized between two operations rather than between operating and not operating.^13,15^ Where the decision under study is whether to undergo surgery at all, rather than which operative technique to use, willingness must be measured in the population of people affected.

Willingness to accept randomization did not meaningfully vary by myelopathy severity. Including patients with moderate or severe disease, for whom surgery is recommended, would be expected to lower willingness to be randomized; that willingness among patients with mild disease was similar suggests the overall estimate was not driven by these patients. Reluctance therefore appears to be a general feature of this treatment decision rather than a function of symptom burden. The reasons patients gave for declining randomization are also informative. Among those unwilling to accept random allocation, 85% cited a wish to maintain control over their treatment decision, whereas fear of surgery was only endorsed by a single respondent and distrust of randomization only endorsed by three respondents. Reluctance arising from fear, or from misunderstanding of trial procedures, is potentially modifiable through education and study procedures.^15^ Reluctance arising from a wish to exercise personal choice is not modifiable in the same way. It is a preference concerning the allocation mechanism itself, which can be accommodated only by altering the study design.

Clinical equipoise, a genuine uncertainty within the expert clinical community as to which of two treatments is preferable, is the ethical precondition for randomization.^25^ Our data are consistent with existing guidelines in suggesting that this condition is met for mild CSM. More than 85% of surgeons considered both surgery and observation reasonable management options, and preference between the two approaches was almost exactly divided, with 46.2% agreeing that early surgery yields better long-term neurological outcomes and 48.1% agreeing that an initial period of observation is more appropriate. More than 80% agreed that further comparative-effectiveness research is required. These attitudes did not differ significantly by specialty training, suggesting that uncertainty is distributed broadly across the surgical community rather than concentrated within a particular subgroup. This finding is a relevant consideration for the feasibility of a broadly generalizable multicenter study, as recruitment would not depend on a small subset of surgeons with atypical views. Surgeon equipoise is, however, necessary but not sufficient: as our patient data show, it does not ensure that patients will accept random allocation.

This distribution of opinion mirrors the state of the published evidence. The 2017 AO Spine/Cervical Spine Research Society guideline was able to recommend only that clinicians offer either surgical intervention or a supervised trial of structured rehabilitation for mild disease, and the systematic review recognizing that recommendation identified no study reporting outcomes exclusively in this population.^7^ Subsequent natural-history data have not converged: reported rates of neurological deterioration under nonoperative management range from 13.3% at one year in a recent cohort restricted to mild disease to 57% over a mean of 2.5 years in an ambispective series.^9,10^ The uncertainty expressed by our respondents is therefore reflective of generally low quality and limited evidence. Equipoise here reflects insufficient evidence rather than demonstrated equivalence.

Overall, these findings, taken with previously published clinical evidence, indicate that the realistic route to comparative evidence in mild CSM is a design that does not require patients to relinquish treatment choice. Well-designed prospective observational studies can yield rigorous comparative effectiveness evidence when treatment preference precludes randomization.^17,18^ The principal limitation of such a design is confounding by indication, and our findings help identify the confounders that would need to be captured. Patients identified worsening neck or arm pain, decline in balance or walking, and deterioration in hand function as the changes most likely to prompt a switch from observation to surgery, rating all three well above decline on clinician-administered testing. These domains would require repeated measurement as time-varying covariates. Patients also identified overall quality of life and physical function as the outcome that mattered most in defining recovery, supporting measurement of broader patient-reported domains such as sleep.^26^ Two-thirds of surgeons reported recommending guideline-concordant structured physical therapy for patients managed nonoperatively, indicating that the appropriate comparator is supervised rehabilitation, often alongside pharmacologic management, rather than an absence of treatment.^7,27^ Surgeons also varied widely in how often they recommended early surgery, from fewer than a quarter to more than half of their patients with mild CSM, so treating surgeon and site should be recorded as potential confounders of treatment selection. Patient factors, such as obesity and physical inactivity, may also influence both treatment selection and outcomes and should likewise be captured.^28^ Pain was the most frequent trigger for considering surgery (64.3%) but was least often selected as the outcome defining recovery (12.5%). Patients therefore appear to treat pain as a marker of progression rather than a treatment goal. Pain should be measured as a time-varying covariate, with function and quality of life as primary endpoints.

### Limitations

Several limitations warrant emphasis. First, and most importantly, we measured stated willingness rather than observed enrollment behavior. Stated willingness does not reliably predict enrollment. In a review of 40 prospective preference assessments, observed randomization rates usually fell between the proportions reporting they were definitely willing and definitely or probably willing, though individual studies both over- and underestimated accrual.^29^ The figure of 28.6% should be treated as an approximate planning estimate rather than a projection. Second, the sample was small and estimates were correspondingly imprecise, particularly within severity strata and in the exploratory analysis of factors associated with willingness. Third, the question regarding randomization preceded that regarding the observational design for all respondents, and an order effect cannot be excluded. However, predominance of autonomy-related reasons for refusal to be randomized argues against this being the principal explanation for the difference in willingness between the two designs. Fourth, 82.7% of responding surgeons practiced in academic settings, and attitudes among community and private-practice surgeons may differ. Surgeon recruitment also used a snowball approach, so an exact response rate could not be calculated and surgeons interested in comparative-effectiveness research may be overrepresented. Most patients (89.3%) identified as White, which limits generalizability to other demographic groups. Finally, while anecdotal refusal rates to participate were low, such events were not prospectively recorded, and therefore potential selection bias cannot be precisely quantified.

## Conclusion

The optimal management of mild CSM remains poorly defined. It has been formally designated as an international research priority,^11,12^ yet the trial that could resolve the question has never been attempted. Our findings indicate that a conventional randomized trial in this population would face difficulty recruiting patients, whereas a prospective observational study appears feasible. Surgeons treating mild CSM are collectively uncertain as to the best management and are highly willing to participate in prospective studies. These findings can help guide the design of a future prospective comparative effectiveness study to inform the benefits and risks of surgery versus nonoperative treatment for mild CSM.

## Data Availability

All data produced in the present study are available upon reasonable request to the authors

