## Supplementary material for "Patient and Surgeon Willingness to Participate in a Randomized Trial of Surgery Versus Observation for Mild Cervical Spondylotic Myelopathy: A Cross-Sectional Survey Study": Tables

**Table 1. Baseline characteristics of survey respondents (patients)**

| **Variable** | **Patients (n = 56)** |
| --- | --- |
| **Demographic** |  |
| Age, y, mean ± SD | 64.1 ± 10.3 |
| Sex assigned at birth, n (%) |  |
| Male | 23 (41.1) |
| Female | 33 (58.9) |
| Race, n (%) |  |
| White | 50 (89.3) |
| Black | 6 (10.7) |
| **Education, n (%)** |  |
| Less than high school | 2 (3.6) |
| High school graduate / GED | 23 (41.1) |
| Bachelor's / college degree | 23 (41.1) |
| Graduate / professional degree | 8 (14.3) |
| **Household income, n (%)** |  |
| < $25,000 | 7 (12.5) |
| $25,000–$49,999 | 8 (14.3) |
| $50,000–$74,999 | 12 (21.4) |
| $75,000–$99,999 | 5 (8.9) |
| ≥ $100,000 | 24 (42.9) |
| **Employment status, n (%)** |  |
| Working full time | 16 (28.6) |
| Working part-time | 6 (10.7) |
| Retired | 26 (46.4) |
| Unemployed | 2 (3.6) |
| Disabled | 6 (10.7) |
| **Modified Japanese Orthopedic Association score** |  |
| mJOA arm/hand function (0–5), mean ± SD | 4.0 ± 1.0 |
| mJOA leg/walking function (0–7), mean ± SD | 4.9 ± 1.4 |
| mJOA hand sensation (0–3), mean ± SD | 2.1 ± 0.7 |
| mJOA bladder function (0–3), mean ± SD | 2.6 ± 0.6 |
| mJOA total (0–18), mean ± SD | 13.6 ± 2.6 |
| **mJOA severity, n (%)** |  |
| Mild (total 15-18) | 24 (42.9) |
| Moderate (total 12-14) | 20 (35.7) |
| Severe (total ≤ 11) | 12 (21.4) |
| **Symptom duration, n (%)** |  |
| < 3 months | 3 (5.4) |
| 3–6 months | 6 (10.7) |
| 7–11 months | 8 (14.3) |
| 1–2 years | 12 (21.4) |
| > 2 years | 27 (48.2) |
| Prior spine surgery, n (%) | 18 (32.1) |
| **Current treatment situation, n (%)** |  |
| Already had surgery | 4 (7.1) |
| Planning to have surgery | 14 (25.0) |
| Currently deciding between surgery and observation | 20 (35.7) |
| Observing symptoms without plans for surgery | 14 (25.0) |
| Have not discussed treatment options with a doctor yet | 4 (7.1) |

*GED = General Educational Development; mJOA = modified Japanese Orthopaedic Association; SD = standard deviation.
mJOA severity was classified as mild (≥ 15), moderate (12–14) or severe (≤ 11). Percentages may not sum to 100 because of rounding.*

**Table 2. Baseline characteristics of survey respondents (surgeons)**

| **Variable** | **Surgeons (n = 52)** |
| --- | --- |
| **Demographic** |  |
| Age, y, mean ± SD | 46.4 ± 9.7 |
| Sex assigned at birth, n (%) |  |
| Male | 48 (92.3) |
| Female | 2 (3.8) |
| Prefer not to say | 2 (3.8) |
| **Race, n (%)** |  |
| White | 31 (59.6) |
| Black | 1 (1.9) |
| Asian | 16 (30.8) |
| Other | 3 (5.8) |
| More than 1 race | 1 (1.9) |
| **Years in practice after training, n (%)** |  |
| 1-5 years | 17 (32.7) |
| 6-10 years | 13 (25.0) |
| 11-15 years | 2 (3.8) |
| 16-20 years | 9 (17.3) |
| More than 20 years | 11 (21.2) |
| **Primary specialty training, n (%)** |  |
| Orthopedic spine surgery | 23 (44.2) |
| Neurosurgery | 24 (46.2) |
| Dual training | 5 (9.6) |
| **Practice environment, n (%)** |  |
| Academic practice | 43 (82.7) |
| Private practice | 4 (7.7) |
| Hospital employed (non-academic) | 3 (5.8) |
| Other | 2 (3.8) |
| **Clinical** |  |
| Mild CSM patients seen per year, n (%) |  |
| Less than 10 | 1 (1.9) |
| 11-25 | 8 (15.4) |
| 26-50 | 21 (40.4) |
| More than 50 | 22 (42.3) |

*CSM = Cervical Spondylotic Myelopathy; SD = standard deviation*.

**Table 3. Univariable associations with willingness to participate in a randomized trial (N = 56; 16 events)**

| **Predictor** | **Univariate logistic OR (95% CI)** |
| --- | --- |
| **Income (per 1-level increase)** | **0.61 (0.39–0.92) p = 0.021** |
| **Deciding between surgery and observation** | **3.39 (1.03–11.78) p = 0.047** |
| Symptom duration (per 1-level increase) | 0.87 (0.55–1.40) p = 0.563 |
| mJOA severity (per 1-level increase) | 1.06 (0.49–2.25) p = 0.870 |
| Sex (female vs male) | 1.80 (0.55–6.60) p = 0.348 |
| Prior spine surgery (yes vs no) | 1.40 (0.40–4.70) p = 0.588 |
| Education (per 1-level increase) | 1.07 (0.49–2.30) p = 0.868 |
| Age (per year) | 1.01 (0.96–1.08) p = 0.645 |

*CI = confidence interval; mJOA = modified Japanese Orthopaedic Association; OR = odds ratio.
Willingness was defined as a response of “likely” or “very likely.” Each predictor was examined in a separate univariable logistic regression model. Income, education, symptom duration and mJOA severity were modeled as ordinal variables (per one-category increase). “Deciding between surgery and observation” compares patients actively deciding with all other treatment-pathway positions.*

**Table 4. Univariable associations with willingness to participate in an observational study (N = 56; 41 events)**

| **Predictor** | **Univariate logistic OR (95% CI)** |
| --- | --- |
| **Sex (female vs male)** | **6.65 (1.76–25.07) p = 0.005** |
| **Deciding between surgery and observation** | **12.09 (1.45–100.69) p = 0.021** |
| Age (per year) | 0.95 (0.90–1.02) p = 0.156 |
| mJOA severity | 0.62 (0.29–1.32) p = 0.215 |
| Education | 1.36 (0.61–3.00) p = 0.452 |
| Symptom duration | 0.68 (0.39–1.20) p = 0.188 |
| Income | 1.10 (0.74–1.64) p = 0.635 |
| Prior spine surgery (yes vs no) | 0.93 (0.26–3.27) p = 0.908 |

*CI = confidence interval; mJOA = modified Japanese Orthopaedic Association; OR = odds ratio.
Willingness was defined as a response of “likely” or “very likely.” Each predictor was examined in a separate univariable logistic regression model. Income, education, symptom duration and mJOA severity were modeled as ordinal variables (per one-category increase). “Deciding between surgery and observation” compares patients actively deciding with all other treatment-pathway positions.*

**Table 5. Surgeon equipoise items (N=52 surgeons)**

| **Variable** | **N (%)** |
| --- | --- |
| **Both surgery and observation are reasonable** |  |
| Disagree | 4 (7.7) |
| Neutral | 3 (5.8) |
| Agree | 27 (51.9) |
| Strongly agree | 18 (34.6) |
| **Early surgery leads to better long-term outcomes** |  |
| Strongly disagree | 2 (3.8) |
| Disagree | 4 (7.7) |
| Neutral | 22 (42.3) |
| Agree | 20 (38.5) |
| Strongly agree | 4 (7.7) |
| **6-12 month observation more appropriate than early surgery** |  |
| Disagree | 11 (21.2) |
| Neutral | 16 (30.8) |
| Agree | 22 (42.3) |
| Strongly agree | 3 (5.8) |
| **Further research is needed** |  |
| Disagree | 3 (5.8) |
| Neutral | 7 (13.5) |
| Agree | 27 (51.9) |
| Strongly agree | 15 (28.8) |

*Response categories selected by no respondents are not shown. Full item wording is provided in Supplementary Data 4.*
