## Supplementary File 3 for "Patient and Surgeon Willingness to Participate in a Randomized Trial of Surgery Versus Observation for Mild Cervical Spondylotic Myelopathy: A Cross-Sectional Survey Study"

**Introductory Information**

You are being asked to complete a short anonymous survey about your views on treatment options for patients with mild symptoms related to spinal cord compression or stenosis. This condition is also known as cervical spondylotic myelopathy (CSM). For patients with mild symptoms, some undergo surgery early on, while others do not and observe their symptoms to see if they worsen. Both treatment options are common and widely accepted. We are trying to understand how we can best design research studies to determine if patients with mild CSM (i.e., mild spinal cord compression) do better with early surgery or non-surgical treatment, like observation and physical therapy. We are hoping you will therefore answer a few questions related to your willingness to participate in a research study addressing this question.

**Treatment Options for Mild CSM**

- **Surgical treatment** aims to relieve pressure on the spinal cord. The goal is to halt further progression and potentially lead to some improvement with symptoms. But there are risks with surgery, such as infection, anesthesia complications, nerve injury, or new pain, among other problems.
- **Non-surgical management** typically includes close observation, sometimes combined with physical therapy. Some people’s symptoms remain stable or possibly improve, while others may worsen over time.

Your answers are completely anonymous.

**SECTION 1: Your Symptoms (mJOA) [Patients Only]**

Please select **ONE** answer in each section.

**1. Which option best describes the function of your arms and hands.**

(Choose one)

0 – Inability to move hands

1 – Inability to eat with a spoon but able to move hands

2 – Inability to button shirt but able to eat with a spoon

3 – Able to button shirt with great difficulty

4 – Able to button shirt with slight difficulty

5 – No dysfunction

**2. Which option best describes the function of your legs and walking ability.** (Choose one)

0 – Complete loss of motor and sensory function

1 – Sensory preservation without ability to move legs

2 – Able to move legs but unable to walk

3 – Able to walk on flat floor with a walking aid (crutch/cane)

4 – Able to walk up/down stairs with handrail

5 – Moderate–significant instability but walk stairs without handrail

6 – Mild instability but walk without aid, smooth reciprocation

7 – No dysfunction

**3. Which option best describes the sensation in your hands.**

0 – Complete loss of hand sensation

1 – Severe sensory loss or pain

2 – Mild sensory loss

3 – No sensory loss

**4. Which option best describes your control over your bladder function?**

(Choose one)

0 – Inability to urinate voluntarily

1 – Marked difficulty with urination

2 – Mild–moderate difficulty with urination

3 – Normal urination

**SECTION 2: Participant Information and Views on Treatment Options**

**5. What is your age?**

(Enter a number)

**6. What is your Sex?**

• Male

• Female

• Prefer not to say

**7. In which U.S. state do you currently live?**

(Select one)

**8. What race do you identify with?**

- White
- Black
- Asian
- Other
- More than 1 race

**9. What is your highest level of education?**

- Less than high school
- High school graduate or GED
- Bachelor’s/college degree
- Graduate or professional degree

10. How would you describe your household income?

- Less than $25,000 per year
- $25,000-$49,999
- $50,000-$74,999
- $75,000-$99,999
- $100,000 or more per year

11. How would you describe your current employment status?

- Working full time
- Working part-time
- Retired
- Unemployed
- Disabled

12. How long have you had symptoms related to spinal cord compression, such as numbness in your arms or legs, clumsiness using your hands, or problems with your walking or balance?

- Less than 3 months
- 3-6 months
- 7-11 months
- 1-2 years
- More than 2 years

13. Have you ever had surgery on any part of your spine before (e.g., neck or lower back), regardless of the reason?

- Yes
- No

**14. Which of the following best describes your current situation regarding treatment for spinal cord compression?**

(Choose one)

- I already had surgery for spinal cord compression
- I am planning to have surgery for spinal cord compression
- I am currently choosing between surgery and observation
- I am currently observing my symptoms without plans to undergo surgery
- I have not discussed treatment options with a doctor yet

**SECTION 3: Your Views on Research Participation**

We are interested in how you feel about participating in a hypothetical research study comparing surgery verssus non-surgical care.

**15. How likely would you be to participate in a research study if you were assigned at random (e.g., with the flip of a coin) to surgery versus observation without surgery for your spinal cord compression?**

- Very unlikely
- Somewhat unlikely
- Neutral
- Likely
- Very likely

**16. If you said you were very unlikely or somewhat unlikely to participate in a randomized study, why would being assigned to a treatment at random be undesirable to you? (select all that apply)**

- Fear of surgery
- Fear or my symptoms worsening if I delay surgery
- I want to maintain control over my treatment decisions
- I don’t trust randomization

**17. How likely would you be to participate in a research study if you were able to choose your own treatment option (i.e., surgery versus observation), based on your discussions with your doctor? The study would involve tracking your symptoms and function over time with whatever treatment option you chose.**

- Very unlikely
- Somewhat unlikely
- Neutral
- Likely
- Very likely

**18. Imagine you were part of a study where you initially pursued or were assigned to non-surgical treatment for your spinal cord compression. What sort of change would make you want to switch from non-surgical treatment to undergoing surgery? (select all that apply)**

- A decline in how I feel my balance and walking abilities are doing.
- Worsening in my ability to use my hands for everyday tasks (e.g., buttoning a shirt)
- Worsening in my neck or arm pain
- Decline in how I performed on tests my doctor gives me, such as my walking speed.

**19. In a study comparing surgery versus non-surgical care for spinal cord compression, what do you think is the single most important outcome to consider when deciding which treatment approach worked better?** (Choose one)

• My overall quality of life and level of physical function

• My ability to perform specific everyday tasks, like walking or ability to button shirts

• My pain (e.g., neck, arm, or shoulder pain)

• Other (please describe): __________________________

**SECTION 4: Optional Feedback**

**20.** **Do you have any comments or concerns about participating in a research study related to the role of surgery for spinal cord compression?**

(Short free-text)
